# Interactive Effects of Maternal Vitamin D Binding Protein and Vitamin D on Offspring Asthma

**DOI:** 10.1101/2025.04.07.25325388

**Authors:** Samuel S. Boyd, Julian Hecker, Hooman Mirzakhani, Dawn L. DeMeo, Scott T. Weiss, Arda Halu

## Abstract

**Rationale:** Vitamin-D-binding-protein transports vitamin-D metabolites and regulates vitamin-D levels in circulation. Additionally, maternal vitamin-D levels during pregnancy plays an important role in lung development and childhood asthma occurrence.

**Objectives:** This study analyzes the joint effect of maternal 25-hydroxyvitamin-D and vitamin-D-binding-protein on offspring asthma.

**Methods:** 806 mother-child pairs who participated in the Vitamin D Antenatal Asthma Reduction Trial were included in this analysis. The primary outcome was offspring asthma by age 3. Maternal plasma vitamin-D-binding-protein levels were measured for 515 participants at 10-18 and 32-38 weeks of gestation. Logistic regression models estimated the relationships between maternal vitamin-D-binding-protein, total 25-hydroxyvitamin-D, and offspring asthma. In addition, offspring asthma was modeled as a function of estimated free 25-hydroxyvitamin-D. A bootstrap approach was used for robust confidence interval estimation.

**Measurements and Main Results:** Maternal vitamin-D-binding-protein levels generally increased as pregnancy progressed. A significant positive interaction effect between maternal vitamin-D-binding-protein and total 25-hydroxyvitamin-D on offspring asthma risk was observed for both the full cohort and the subset of mothers with asthma, suggesting that the protective effect of total 25-hydroxyvitamin-D increases with lower levels of vitamin-D-binding-protein. For mothers with asthma, estimated maternal free 25-hydroxyvitamin-D was found to have a significant protective effect against offspring asthma, surpassing the effects of vitamin-D-binding-protein or total 25-hydroxyvitamin-D individually.

**Conclusions:** These results highlight the interplay between vitamin-D metabolites during pregnancy and their protective effects for offspring asthma. These results also provide evidence for the free hormone hypothesis, which suggests that free vitamin-D is more biologically relevant than total vitamin-D.

## Introduction

Asthma is the second most prevalent chronic respiratory disease, affecting approximately 3.6% of the global population (1). Notably, asthma is the most common chronic respiratory disease among children, with an estimated 14% of children worldwide affected (1). The risk of developing disease in childhood, adolescence, and even adulthood is heavily influenced by prenatal perturbations (2). This is especially true of asthma, where factors such as maternal asthma can result in increased risk of offspring asthma (3).

Vitamin D plays critical roles for healthy fetal development during pregnancy, including maternal immune system modulation, fetal bone development, and fetal lung development (4). Yet there is widespread vitamin D deficiency especially during pregnancy, determined by the circulating levels of 25-hydroxyvitamin D_3_ (25OHD, our primary vitamin D metabolite) (5). The Vitamin D Antenatal Asthma Reduction Trial (VDAART) was designed to investigate whether vitamin D supplementation (4,400 IU vs. 400 IU daily) early in pregnancy (10-18 weeks of gestation) would decrease the risk of offspring asthma and other adverse outcomes such as preeclampsia (6). Although initial findings were statistically insignificant (*OR* = 0.73; 95% *CI* = [0.53, 1.00]) (7), a meta-analysis controlling for baseline 25OHD levels revealed a statistically significant result (*OR* = 0.43; 95% *CI* = [0.19, 0.96]) (8, 9). Even though VDAART and other clinical trials have not shown conclusive evidence of the protective effect of vitamin D on childhood asthma (10), their findings have pointed towards its potential preventative role. Therefore, additional data on other molecules involved in vitamin D metabolism and signaling, such as vitamin D binding protein, may lead to a clearer understanding of vitamin D’s role in the development of childhood asthma.

Vitamin D binding protein (DBP) is a protein encoded by the Group-Specific Component (*GC*) gene that serves to transport various vitamin D metabolites throughout the body, primarily 25OHD (11). Approximately 85% of circulating vitamin D metabolites are bound, primarily to DBP, but also to albumin with much lower affinity. Bound vitamin D metabolites have a longer half-life in circulation (12). DBP thus acts as a circulating reservoir of vitamin D metabolites, including 25OHD. Given DBPs importance in vitamin D metabolism, this study aims to investigate the role that DBP may have in modulating the effect of 25OHD levels on offspring asthma risk. Furthermore, in line with the free hormone hypothesis, which states that it is the free, rather than bound, molecule that is biologically active, we investigate the relationship between estimated free 25OHD levels and offspring asthma incidence.

## Methods

### VDAART Design and Participants

VDAART is a randomized, double blind, placebo-controlled clinical trial of vitamin D supplementation (4,000 IU plus a multivitamin with 400 IU daily) versus placebo (placebo pill plus a multivitamin with 400 IU daily) in pregnant women, given after baseline measurements were taken, to prevent adverse pregnancy outcomes and “asthma and recurrent wheeze” in their children. Eligible participants were women between the ages of 18 and 39 years with the estimated gestational ages between 10 and 18 weeks (singleton), no history of smoking or current nicotine product use, a history of physician-diagnosed asthma or atopy, or a partner (biologic father of the child) with a history of physician-diagnosed asthma or atopy. Eligible participants were screened per VDAART protocol and enrolled if eligible. Details of the trial design, protocol, and primary and secondary outcomes have been published (6, 7, 13). In this study, we included pregnant women enrolled in VDAART who provided any postnatal data, up to the child’s age of 3 years.

### Primary Outcomes

In this study, the primary outcome was offspring asthma incidence by the age of 3 years, defined by a parental report of a physician’s diagnosis of asthma on the prospective study questionnaires. In statistical modeling, absence of offspring asthma by the age of 3 years was treated as the referent group.

### Measurement of Vitamin D Binding Protein

The levels of vitamin D binding protein (DBP) were measured for maternal plasma samples taken at the first and third trimester of pregnancy (10-18 weeks and 32-38 weeks, denoted by M1 and M2, respectively), where baseline first trimester measurements (M1) were taken before administering the intervention. DBP levels were measured by Heartland Assays, Inc. (Ames, Iowa) using R&D Systems Human Vitamin D Binding Protein Quantikine ELISA Kit (R&D Systems, Minneapolis, Minnesota), which employs the quantitative sandwich enzyme immunoassay technique (14).

### Estimation of Free Vitamin D Levels

The free hormone hypothesis postulates that the biological activity of a biomolecule depends primarily on its unbound concentration, and there is existing evidence that this applies to vitamin D metabolites (12). Due to the absence of directly measured free 25OHD levels, estimates of these levels were derived from total 25OHD and DBP levels, following similar calculations in (12). Specifically, free 25OHD levels were estimated by the ratio of total 25OHD to DBP levels.

### Additional Variables

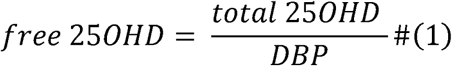

Several additional covariates were included in the statistical modeling that are known to affect offspring asthma development and other variables of interest such as 25OHD and DBP levels. Prenatal covariates included maternal age, maternal asthma status, maternal race/ethnicity, maternal baseline BMI, clinical site, and treatment group. Additionally, season of vitamin D measurement and gestational age at DBP measurement were included in models with 25OHD and DBP, respectively. Post-partum covariates included gestational age at delivery and offspring sex. For outcomes coming from post-partum samples (*i.e.*, cord blood, offspring), both the pre-partum and post-partum covariate sets were included in the model. Additionally, the relative change from the first to third trimester (M1 to M2) for 25OHD (free and total) and DBP were calculated as 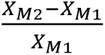 and investigated for associations with offspring asthma incidence.

### Statistical Analysis

For comparing differences in distribution of various clinical and demographic variables between mothers whose children did versus did not develop asthma by the age of 3 years, we performed Mann-Whitney-Wilcoxon tests for continuous variables and Pearson’s χ2 tests for categorical variables. Logistic regression models of offspring asthma incidence were estimated. Bootstrapped confidence intervals (bCI) were obtained for model coefficients by taking 10,000 samples drawn with replacement from the study cohort, fitting models to these bootstrapped samples, and obtaining a background distribution of coefficient estimates. The 95% bCI ranged from the 2.5th percentile to the 97.5th percentile of the background distribution. A coefficient estimate was considered statistically significant if its 95% bCI excluded zero. In addition to the full cohort analysis, we analyzed the subset of mothers with asthma, with maternal asthma being a known risk factor for offspring asthma (15). The family-wise error rate (FWER) and false discovery rate (FDR) were not controlled for. A Type I error rate of 0.05 was used for all statistical analyses in this study. R version 4.2.0 was used for all analyses (R Foundation for Statistical Computing).

## Results

### Study Cohort Characteristics

Of the 806 VDAART participants included in this study, 125 gave birth to children who developed asthma by 3 years (**Table 1**). First and third trimester total 25OHD levels are significantly lower in mothers of children who developed asthma by age 3, compared to those whose children did not develop asthma (*p_M_*_l_ = 2.0 × 10^−4^; *p_M_*_2_ = 1.0× 10^−3^). One can see analogous results for the comparison of estimated free 25OHD levels by offspring asthma status.

**Table 1.** Characteristics of study subjects stratified by offspring asthma status by the age of three years. Differences between groups were assessed using Mann-Whitney-Wilcoxon tests and Pearson’s x^2^ tests for continuous and categorical variables, respectively. Asterisks represent statistically significant differences, with a Type I error rate controlled at 0.05.

|  | Offspring Asthma (by age 3) |  |  |
| --- | --- | --- | --- |
|  | No<br>(N=681) | Yes<br>(N=125) | Total<br>(N=806) |
| **Total 25OHD (ng/mL)(M1) \*** |  |  |  |
| Mean (SD) | 23.5 (10.4) | 19.8 (8.83) | 23.0 (10.2) |
| Missing | 3 (0.4%) | 2 (1.6%) | 5 (0.6%) |
| **Total 25OHD (ng/mL)(M2) \*** |  |  |  |
| Mean (SD) | 33.7 (14.4) | 29.2 (15.3) | 33.0 (14.6) |
| Missing | 28 (4.1%) | 9 (7.2%) | 37 (4.6%) |
| **Relative Change in Total 25OHD (M1 to M2)** |  |  |  |
| Mean (SD) | 0.625 (0.911) | 0.627 (1.01) | 0.625 (0.925) |
| Missing | 30 (4.4%) | 11 (8.8%) | 41 (5.1%) |
|  | No<br>(N=681) | Yes<br>(N=125) | Total<br>(N=806) |
| **DBP (µg/mL)(M1)** |  |  |  |
| Mean (SD) | 517 (128) | 534 (134) | 520 (129) |
| Missing | 246 (36.1%) | 43 (34.4%) | 289 (35.9%) |
| **DBP (µg/mL)(M2)** |  |  |  |
| Mean (SD) | 667 (200) | 691 (233) | 670 (206) |
| Missing | 246 (36.1%) | 44 (35.2%) | 290 (36.0%) |
| **Relative Change in DBP (M1 to M2)** |  |  |  |
| Mean (SD) | 0.328 (0.391) | 0.334 (0.440) | 0.329 (0.398) |
| Missing | 247 (36.3%) | 44 (35.2%) | 291 (36.1%) |
| **Estimated Free 25OHD (M1) \*** |  |  |  |
| Mean (SD) | 0.0500 (0.0258) | 0.0406 (0.0179) | 0.0485 (0.0249) |
| Missing | 247 (36.3%) | 43 (34.4%) | 290 (36.0%) |
| **Estimated Free 25OHD (M2) \*** |  |  |  |
| Mean (SD) | 0.0555 (0.0278) | 0.0462 (0.0266) | 0.0541 (0.0278) |
| Missing | 247 (36.3%) | 44 (35.2%) | 291 (36.1%) |
| **Relative Change in Free 25OHD (M1 to M2)** |  |  |  |
| Mean (SD) | 0.307 (0.861) | 0.273 (0.771) | 0.301 (0.847) |
| Missing | 249 (36.6%) | 44 (35.2%) | 293 (36.4%) |
| **Maternal Age \*** |  |  |  |
| Mean (SD) | 27.7 (5.43) | 25.5 (5.58) | 27.3 (5.51) |
| **Maternal BMI (M1) \*** |  |  |  |
| Mean (SD) | 28.5 (7.50) | 31.3 (8.08) | 28.9 (7.65) |
| Missing | 36 (5.3%) | 7 (5.6%) | 43 (5.3%) |
| **Maternal Asthma Status \*** |  |  |  |

| Offspring Asthma (by age 3) |  |  |  |
| --- | --- | --- | --- |
|  | No<br>(N=681) | Yes<br>(N=125) | Total<br>(N=806) |
| No | 433 (63.6%) | 51 (40.8%) | 484 (60.0%) |
| Yes | 248 (36.4%) | 74 (59.2%) | 322 (40.0%) |
| **Maternal Race/Ethnicity \*** |  |  |  |
| Caucasian (Non-Hispanic) | 190 (27.9%) | 24 (19.2%) | 214 (26.6%) |
| African American (Hispanic and Non-Hispanic) | 270 (39.6%) | 77 (61.6%) | 347 (43.1%) |
| Caucasian (Hispanic) | 95 (14.0%) | 17 (13.6%) | 112 (13.9%) |
| Other | 108 (15.9%) | 4 (3.2%) | 112 (13.9%) |
| Missing | 18 (2.6%) | 3 (2.4%) | 21 (2.6%) |
| **Site \*** |  |  |  |
| Boston | 201 (29.5%) | 39 (31.2%) | 240 (29.8%) |
| San Diego | 250 (36.7%) | 24 (19.2%) | 274 (34.0%) |
| St. Louis | 230 (33.8%) | 62 (49.6%) | 292 (36.2%) |
| **Treatment** |  |  |  |
| Vitamin D Supplementation | 336 (49.3%) | 65 (52.0%) | 401 (49.8%) |
| Placebo | 345 (50.7%) | 60 (48.0%) | 405 (50.2%) |
| **Child Sex \*** |  |  |  |
| Female | 339 (49.8%) | 46 (36.8%) | 385 (47.8%) |
| Male | 342 (50.2%) | 79 (63.2%) | 421 (52.2%) |

The maternal asthma rate for the mothers of children who developed asthma is 59.2%, compared to a rate of 36.4% for mothers of children who did not develop asthma (*p* = 2.9 × 10^−6^). There is a significantly higher rate of male offspring among those who developed asthma (*p* = 0.01). Additionally, mothers of children who developed asthma have a significantly higher baseline BMI (*p* = 3.3 × 10^−4^). This difference in BMI is strengthened (in terms of a lower p-value) when considering the subset of mothers with asthma, and this difference vanishes when considering the subset of mothers that do not have asthma (**Figure E1** in the online data supplement).

### Maternal DBP Levels Increase During Pregnancy

To explore the impact that DBP may have on offspring asthma risk, DBP levels were measured for 517 and 516 maternal blood samples taken at the first (M1, pre-intervention) and third (M2, post-intervention) trimesters, respectively, with 515 participants having samples at both timepoints (**Table 1**). Maternal DBP levels do not differ significantly by offspring asthma status or treatment group. Mean DBP levels increased from 520 μg/mL to 670 μg/mL between the first and third trimesters. 85% of mothers showed an increase in DBP levels, with an average relative change of 41%, while only 15% of mothers showed a slight decrease in DBP (**Figure 1**). The sign of the DBP change between trimesters was not significantly associated with vitamin D levels or treatment group.

**Figure 1.**
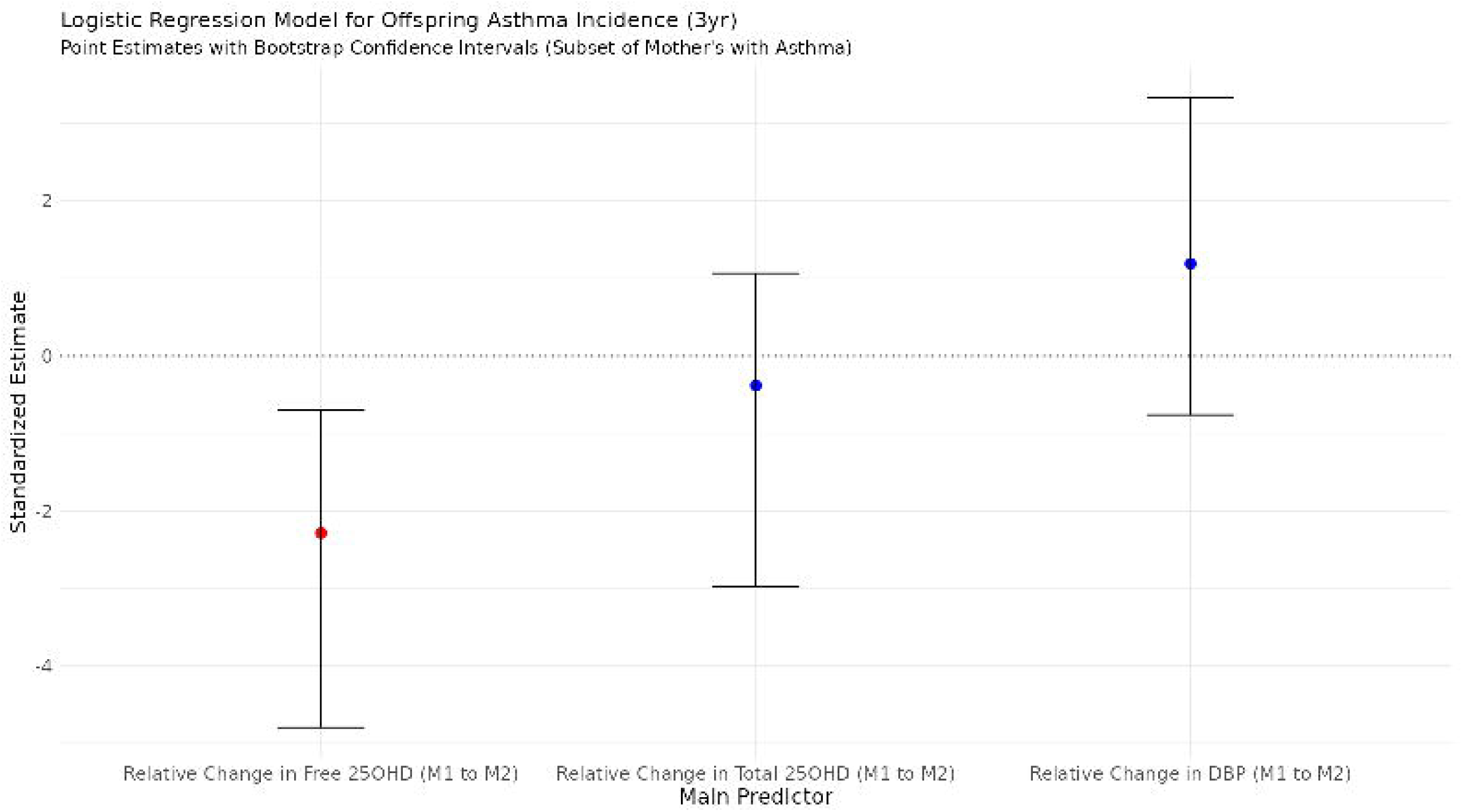
Maternal DBP Levels During Pregnancy. Red (blue) indicates an increase (decrease) in DBP levels from the first to third trimester (M1 to M2). Percentages indicate the percentage of participants with an increase or decrease. Avg. rel. change = average relative change from M1 to M2.

### Maternal DBP and 25OHD Levels Jointly Affect Offspring Asthma Risk

To investigate whether maternal DBP levels modulate the effect of maternal total 25OHD levels on offspring asthma risk, we estimated a logistic regression model of offspring asthma incidence, which included an interaction term between DBP and 25OHD, along with possibly confounding variables, including treatment group and maternal asthma status (see Methods). For the full cohort, we fit two models for baseline (M1) and third trimester (M2) measurements of DBP and 25OHD. The interaction effect between DBP and 25OHD on offspring asthma was statistically significant for the first trimester model only (*β* = 2.11 × 10^−4^; bCI = [3.9 × 10^−5^, 4.54 × 10^−4^]). Conditional main effects of DBP and 25OHD levels on offspring asthma risk can be seen in **Figure 2**. For mothers with lower baseline DBP levels, the protective effect of 25OHD against childhood asthma was strengthened, with this conditional main effect of vitamin D being statistically significant for low levels of DBP (**Figure 2, right**). On the other hand, there was a statistically significant adverse effect of DBP on asthma risk for higher levels of 25OHD (**Figure 2, left**).

**Figure 2.**
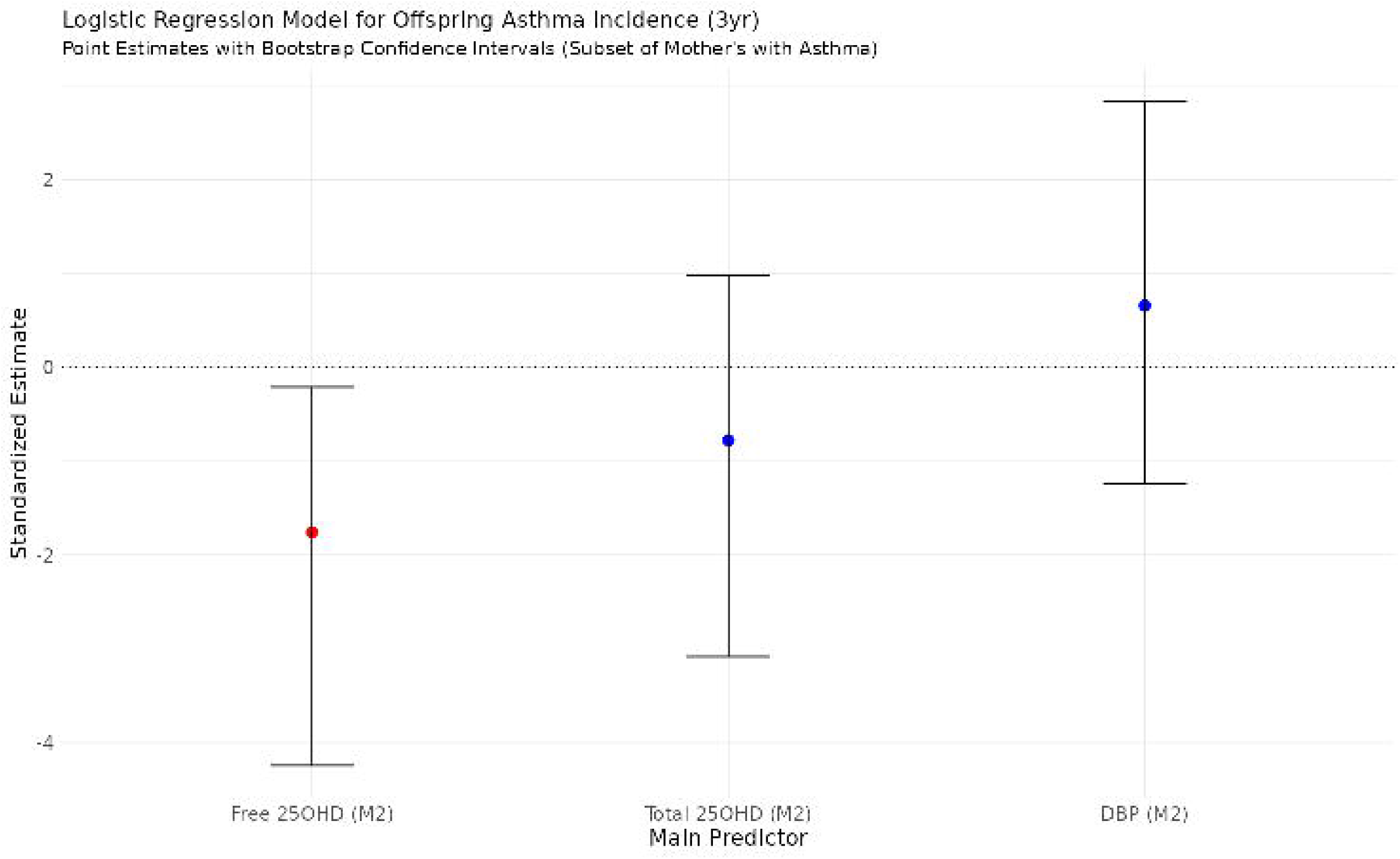
Conditional Main Effects of 25OHD x DBP Interaction Model for Offspring Asthma at 3 years (Full cohort). Conditional main effects and standard errors were estimated by centering the conditioning variable (i.e., variable on x-axis) by a grid of values within the observed range and refitting the model. These estimates and standard errors were then used to assess statistical significance of the conditioned variable effect (i.e., y-axis) for each value of the conditioning variable. Bars at top of figure show histograms of the x-axis variable for the full VDAART cohort. Left and right plots have the same y-axis scale.

This analysis was repeated for the subset of mothers with asthma (N=322) to investigate the potential 25OHD-DBP interaction effect for this at-risk population. Similar to the previous findings, the interaction effect between DBP and 25OHD on offspring asthma was statistically significant for the baseline model only (*β* = 4.21 × 10^−4^; bCI = [1.54 × 10^−4^, 8.66 × 10^−4^]). Conditional main effects of DBP and 25OHD levels on offspring asthma risk, for the subset of mothers with asthma, can be seen in **Figure 3**. There is an increased protective effect of baseline 25OHD against offspring asthma for lower levels of baseline DBP (**Figure 3, right**). Furthermore, our data show an increased protective effect of DBP for low baseline levels of 25OHD, as well as an adverse effect of DBP for higher baseline levels of 25OHD (**Figure 3, left**). These results did not hold for the subset of mothers without asthma.

**Figure 3.**
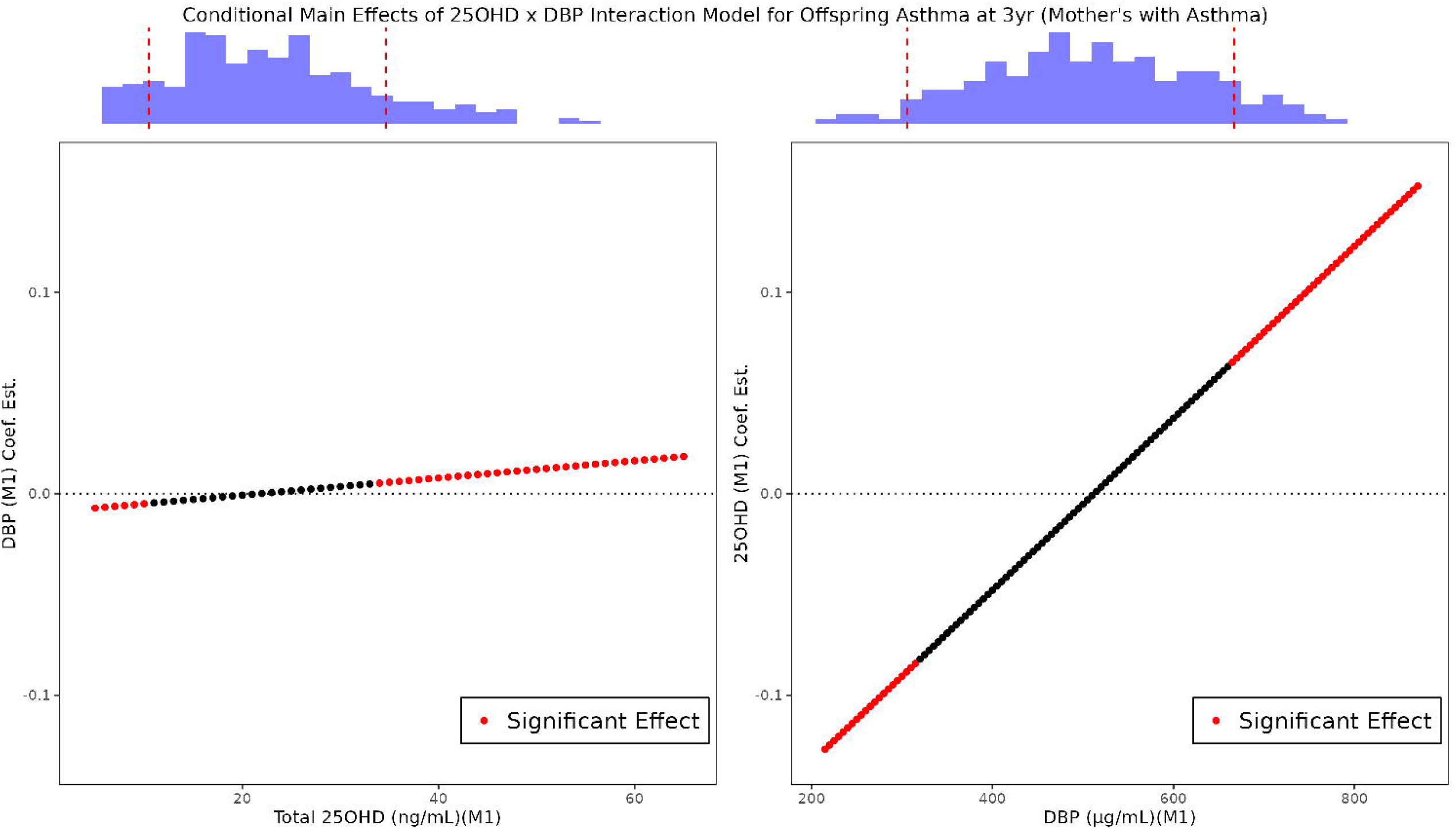
Conditional Main Effects of 25OHD x DBP Interaction Model for Offspring Asthma at 3 years (Mothers with asthma). Conditional main effects and standard errors were estimated by centering the conditioning variable (i.e., variable on x-axis) by a grid of values within the observed range and refitting the model. These estimates and standard errors were then used to assess statistical significance of the conditioned variable effect (i.e., y-axis) for each value of the conditioning variable. Bars at top of figure show histograms of the x-axis variable for the subset of VDAART mothers with asthma. Left and right plots have the same y-axis scale.

### Estimated Maternal Free 25OHD Levels Are More Predictive of Offspring Asthma than Total 25OHD

It is hypothesized that the free, rather than bound, levels of 25OHD are more biologically relevant. Due to the absence of directly measured free 25OHD levels, these were estimated using existing maternal DBP and total 25OHD measurements (Methods). For the subset of mothers with asthma, logistic regression models of offspring asthma incidence were fit with either free 25OHD, total 25OHD, or DBP levels as the main predictor. While total 25OHD and DBP levels at the third trimester (M2) were not statistically significant, estimated free 25OHD showed a significant protective effect against offspring asthma (**Figure 4**). Furthermore, this result holds when considering relative change in estimated free 25OHD from M1 to M2 (**Figure 5**). These results were not replicated in either the full cohort or the subset of mothers without asthma.

**Figure 4.**
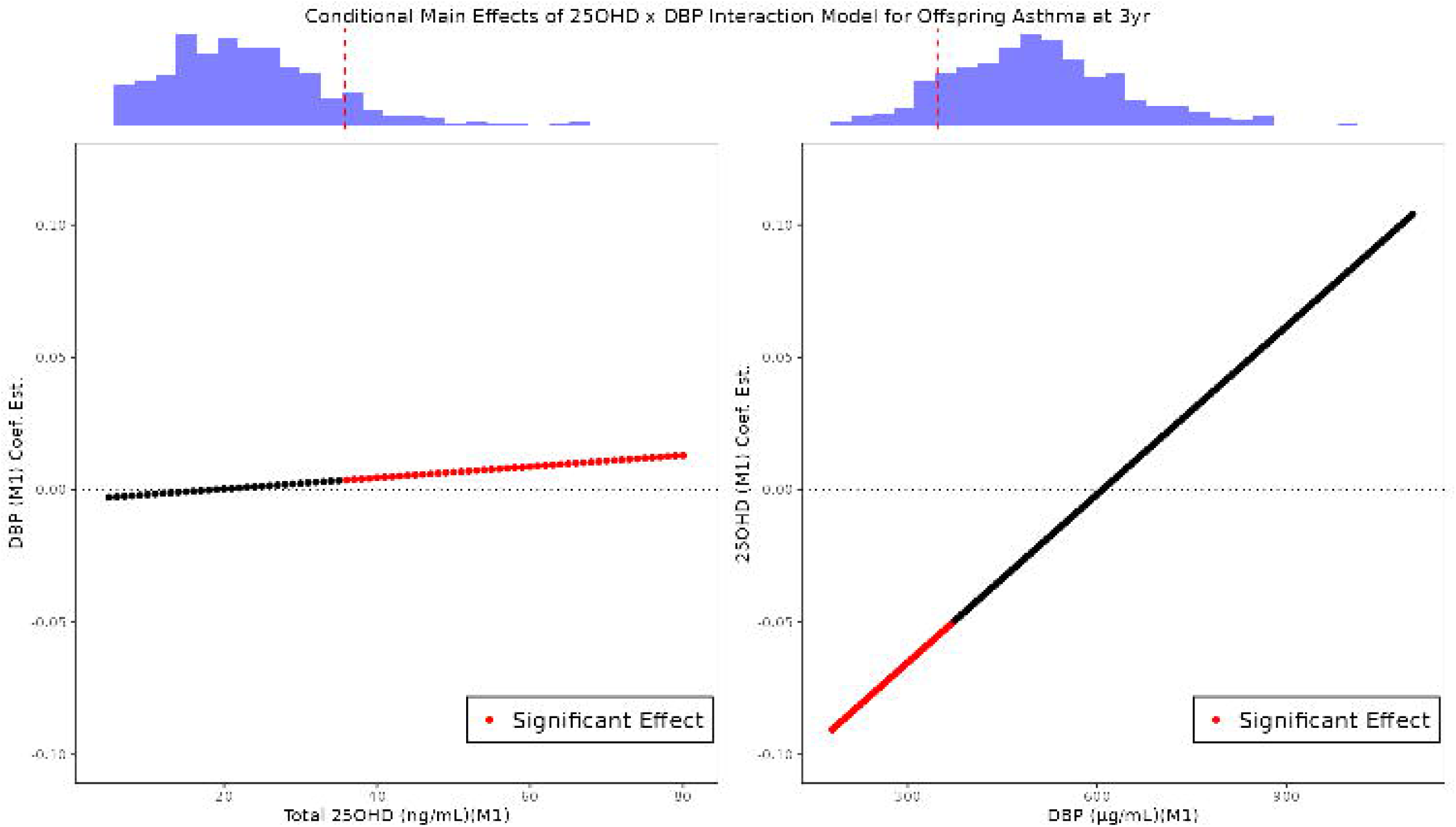
Point Estimates with Bootstrapped Confidence Intervals for Logistic Regression Models of Offspring Asthma (Full cohort). Confidence Intervals (CI) are found by taking the 2.5^th^ and 97.5^th^ percentile of the bootstrap distribution of the coefficient estimates. 10,000 bootstrap samples were used. Point estimates and bootstrapped CIs were scaled by the standard deviation of the bootstrap distribution.

**Figure 5.**
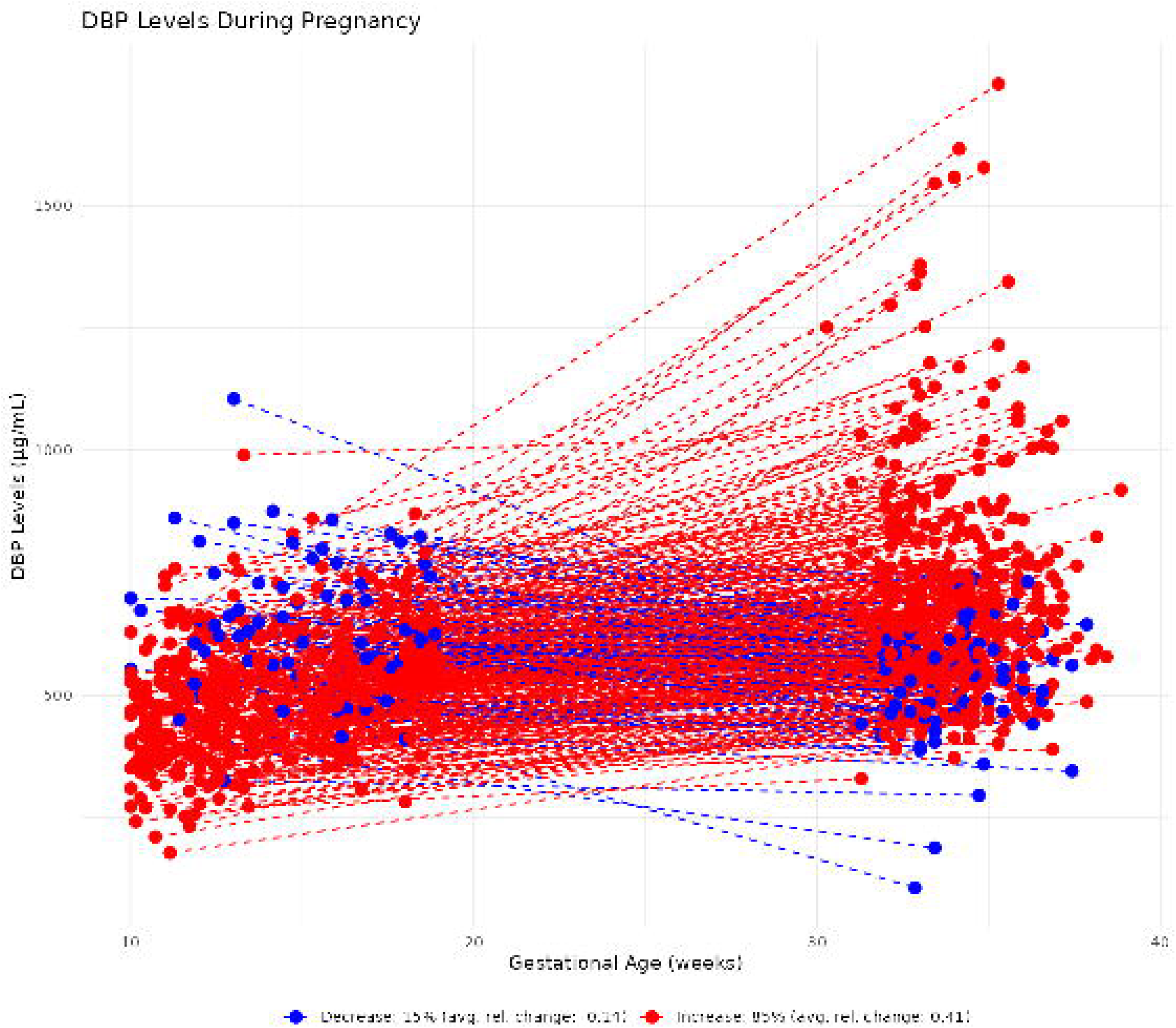
Point Estimates with Bootstrapped Confidence Intervals for Logistic Regression Models of Offspring Asthma (Mothers with asthma). Confidence Intervals (CI) are found by taking the 2.5^th^ and 97.5^th^ percentile of the bootstrap distribution of the coefficient estimates. 10,000 bootstrap samples were used. Point estimates and bootstrapped CIs were scaled by the standard deviation of the bootstrap distribution.

## Discussion

This study focused on newly measured maternal DBP levels of plasma samples from VDAART participants. We found a general increase in DBP levels from the first to third trimester, although there is a small proportion of participants showing a slight decrease in DBP. We investigated the interactive effect between maternal total 25OHD and DBP on offspring asthma risk by the age of 3 years. For both the full cohort and the subset of mothers with asthma, this interaction effect was statistically significant. We also identified estimated free 25OHD as more predictive of offspring asthma incidence than either total 25OHD or DBP individually.

Maternal DBP levels did not differ significantly between treatment groups. However, DBP levels did differ between timepoints. We observed a general increase in DBP levels from the first to third trimester, although 15% of mothers showed a relatively small decrease. It is hypothesized that the measured increase or decrease in DBP levels is primarily due to the gestational age at measurements. DBP levels naturally increase during pregnancy, likely due to increased estrogen levels (16). During pregnancy, DBP concentrations reach a maximum around week 28 of gestation (17). Furthermore, mothers with an observed decrease in DBP had significantly higher DBP levels and a later gestational age at M1 (**Figure E2** in the online data supplement). Therefore, the variability in DBP increase vs. decrease is most likely due to the timing of an individual’s peak with respect to their gestational age at measurements M1 and M2.

In the DBPxTotal-25OHD interaction model of offspring asthma, the main effect of total 25OHD is conditional on DBP, and vice versa. The protective effect of total 25OHD levels on childhood asthma incidence by 3 years increases as DBP levels decrease, with this conditional effect being statistically significant for DBP levels below 370 µg/mL for the full cohort (below 315 µg/mL for the subset of mothers with asthma). Additionally, there is an increasing adverse effect of DBP on offspring asthma risk as total 25OHD levels increase, with this conditional effect being statistically significant with total 25OHD levels above 36 ng/mL for the full cohort (above 34 ng/mL for the subset of mothers with asthma). However, the magnitude of the total 25OHD conditional main effect is almost an order of magnitude larger than that of DBP at the extremes. This finding suggests that vitamin D supplementation may be most beneficial, in terms of asthma risk reduction, to pregnant women with lower levels of DBP.

We next investigated the association of estimated free 25OHD levels with offspring asthma incidence. This was estimated. In our analysis, free 25OHD was approximated as the ratio of total 25OHD levels to DBP levels. Out of necessity, this is a more simplistic estimation than in (12), which considers albumin levels and binding affinities. Albumin measurements did not exist for our samples; furthermore, 25OHD-to-DBP binding affinities are highly variable between *GC* genotypes and populations (12). For the subset of mothers with asthma, we found that higher levels of free 25OHD in the third trimester, along with larger relative changes in free 25OHD from the first to third trimesters, are significantly associated with reduced offspring asthma risk. These models included treatment group as a covariate. Importantly, these effects were stronger than the individual effects of total 25OHD or DBP. In light of the results from the interaction model, these significant results for free 25OHD may not be surprising. The significant interaction effect between total 25OHD and DBP indicates that their joint effect is non-additive, meaning the effect of one variable depends on the level of the other. Since our free 25OHD levels are estimated by the ratio of total 25OHD to DBP, this may be capturing part of this non-additive relationship between total 25OHD, DBP, and asthma incidence.

A primary limitation of this study is the lack of DBP measurements in potential replication cohorts. Thus, we currently do not know if these results generalize to other populations. Another limitation is the absence of directly measured free vitamin D. As mentioned previously, our estimation of free vitamin D is necessarily simpler than other estimation methods, although it has been shown that these more sophisticated methods still show weak correlation with directly measured free vitamin D (18). Additionally, this study explored multiple associations. Given the exploratory nature of this work and the small number of hypotheses tested, corrections for multiple testing (to control FWER or FDR) were not applied, as this could reduce statistical power and increase the risk of Type II errors (19). Findings should be interpreted with caution, and future confirmatory studies are needed to validate these associations.

In conclusion, we found a significantly positive interaction effect between DBP and total 25OHD on offspring asthma incidence, for both the full cohort and the subset of mothers with asthma. Additionally, our free 25OHD estimates show a stronger association with offspring asthma incidence than total 25OHD or DBP individually, especially when mothers have a history of asthma. These observations highlight that DBP and free vitamin D can provide important missing links for explaining the relationship between maternal vitamin D and offspring asthma.

## Data Availability

Data is available upon request.

**Figure E1.** Baseline BMI Stratified by Offspring Asthma Status. Boxplots comparing distribution of maternal baseline BMI by offspring asthma status. P-values come from a non-parametric Mann-Whitney-Wilcoxon test.

**Figure E2.** Maternal DBP levels and Gestational Ages at M1 by DBP Increase/Decrease Status. Boxplots comparing distributions of maternal DBP levels and gestational age at M1 by DBP Increase vs. Decrease status (regarding the change from M1 to M2). P-values come from a non-parametric Mann-Whitney-Wilcoxon test.

