## Supplementary figures and images for "Interactive Effects of Maternal Vitamin D Binding Protein and Vitamin D on Offspring Asthma"

### FigureE1.png

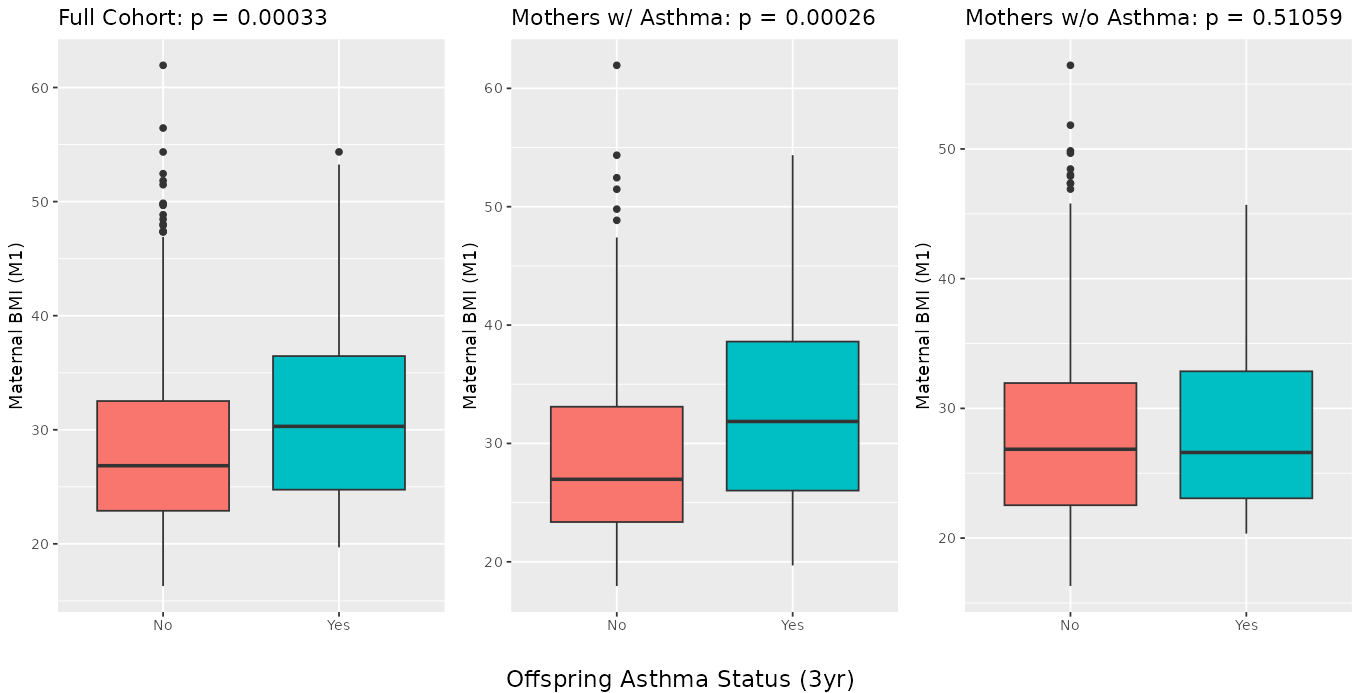

### FigureE2.png

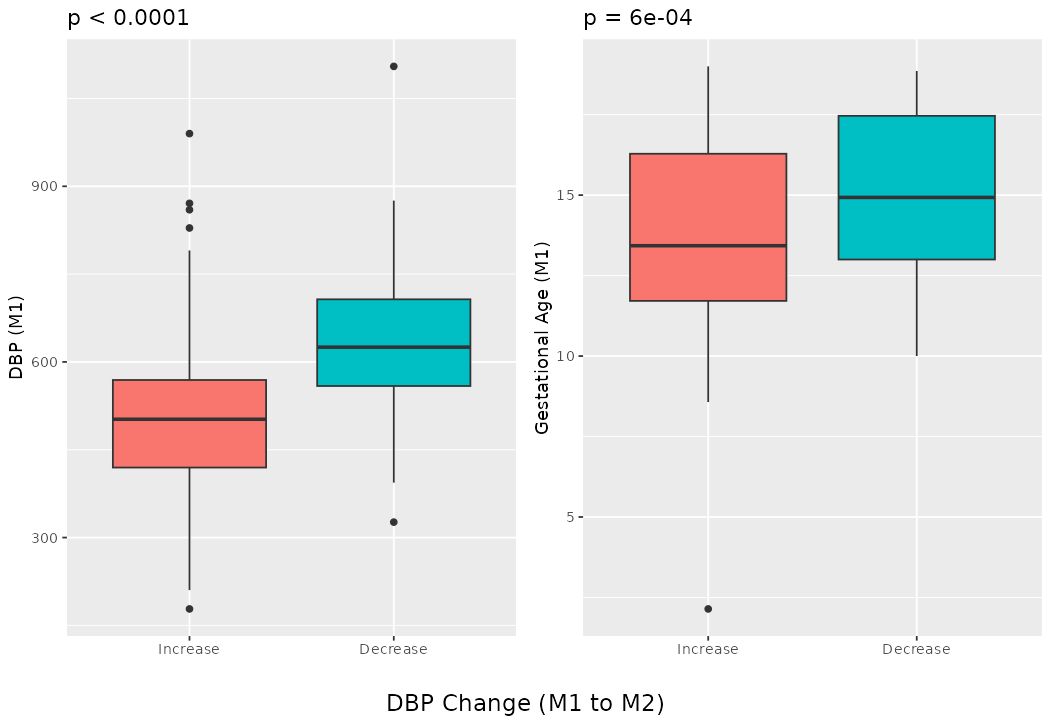
